# Radiobiological analysis of the response of prostate cancer to different fractionations

**DOI:** 10.1101/2023.02.28.23286507

**Authors:** Juan Pardo-Montero, Isabel González-Crespo, Antonio Gómez-Caamaño, Araceli Gago-Arias

## Abstract

**Purpose:** To investigate the response of prostate cancer to different radiotherapy schedules, including hypofractionation, and to evaluate potential departures from the linear-quadratic (LQ) response. To obtain best-fitting parameters for low (LR), intermediate (IR), and high risk (HR) prostate cancer.

**Methods and Materials:** We have constructed a dataset of dose-response containing 87 entries (35 LR, 32 IR, 20 HR), with doses per fraction ranging from 1.8 to 10 Gy. These data were fitted to tumor control probability models based on the LQ model, linear-quadratic-linear (LQL), and a modification of the LQ (LQmod) accounting for increasing radiosensitivity at large doses. Fits were performed with the maximum likelihood expectation methodology, and the Akaike-Information-Criterion (AIC) was used to compare models.

**Results:** The AIC shows that the LQ model is superior to the LQL and LQmod for all risks, except for IR where the LQL outperforms the other models. The analysis shows a low α/β for all risks: 2.01 Gy for LR (95% confidence interval 1.74-2.26), 3.44 Gy for IR (2.99-4.02), and 2.78 Gy for HR (1.43-4.18). Best-fits do not show proliferation for LR, and only moderate proliferation for IR/HR.

**Conclusions:** In general, the LQ model describes the response of prostate cancer better than the alternative models. Only for IR the LQL outperforms the LQ. This study confirms a low *α/β* for all risks, with doses per fraction ranging from <2 Gy up to 10 Gy.

## Introduction

The response of prostate cancer to radiotherapy has been extensively analyzed in the radiobiological modeling literature **[1-10]**. Most studies report a low *α/β* (typically in the 1-3 Gy range) and high sensitivity to fractionation, even though some studies suggest that the *α/β* may not be that low and the reported low values may be caused by hypoxia **[8]**.

A low *α/β* for prostate cancer, lower than the *α/β* associated with late toxicities of nearby tissues, may favour hypofractionated regimes. In recent years, Stereotactic Body RadioTherapy (SBRT) has become widely used to treat many cancers **[11]**, and several trials have explored the response/toxicity of hypofractionation in prostate cancer **[12-14]**, with doses per fraction reaching up to 10 Gy.

The validity of the linear-quadratic (LQ) model for large dose fractions has been questioned **[15-17]**. Some studies point out a moderation of the LQ cell killing effect with increasing dose, an effect that has been modeled with the linear-quadratic-linear (LQL) and other approaches **[18, 19]**. Also, recent *in vivo* studies have shown an enhanced cell killing effect at large doses attributed to *indirect* effects like vascular damage and radiation-induced immune-response **[20-22]**, which has led to novel models including such effects **[23-26]**.

Because the implementation of hypofractionation for prostate cancer is relatively new, there are not many radiobiological modeling studies investigating the response of prostate cancer to hypofractionation. We have to notice the study by Datta *et al*. **[10]**, who analyzed 8 isoeffective schedules (conventional and hypofractionated) and obtained a *α/β* value in the 1.3-11.1 Gy range. Also, a recent study by Royce *et al*. **[27]** analyzed the tumor control probability (TCP) of 25 hypofractionated clinical studies and obtained the EQD2 needed to reach 90-95% control by assuming *α/β* =1.5 Gy.

In this work we will further explore the radiobiology of prostate cancer with a large dataset of treatments, with doses-per-fraction ranging from <2 Gy to 10 Gy. Our aim is two-fold: on the one hand, we will evaluate whether the addition of dose-response data for severely hypofractionated schedules leads to deviations from the LQ model, by comparing best-fits obtained with the LQ model and other models. On the other hand, we will determine best-fitting radiobiological parameters that describe the response of prostate cancer to fractionation, split by risk level, in a large dataset containing a wide range of fractionations.

## Materials and methods

### Clinical dataset

We have analyzed dose-response data for 55 trials of prostate radiotherapy, building on data previously compiled in several radiobiological studies by Royce, Miralbell, Datta and Pedicini and colleagues **[4, 9, 10, 27]**. The original references were reviewed to avoid any inconsistency. For each schedule, we extracted the number of patients, distribution of patients for risk level, number or percentange of patients receiving androgen deprivation therapy (ADT), dose per fraction, total dose, treatment time, and control at 5 years. Some studies included slightly different fractionations, and in those cases the most used fractionation was included. Control may be named differently in publications, but it generally refers to *freedom from biochemical failure*, with biochemical failure defined as PSA nadir + 2 ng/mL. We have restricted our analysis to studies reporting control at 5 years: prostate cancer is usually a slow-growing disease and differences in control between different schedules may not be significant at 3 years. On the other hand, some studies also reported control at 7-7.5 years, but those data were discarded because there were very few of them. Control values were generally reported in the text, but sometimes were extracted from figures by using image analysis software *(g3data)*.

When separating by risk, we analyzed 35, 32 and 20 schedules for low risk (LR), intermediate risk (IR) and high risk (HR), respectively. Some studies included extra groups, like “favorable intermediate risk”, “unfavorable intermediate risk” and “very low risk”. In such cases, those results were merged in a single group (favorable and unfavorable intermediate risk merged in “intermediate risk”; very low risk and low risk merged in “low risk”).

Many of the clinical protocols included androgen deprivation therapy (ADT). In general, LR patients did not receive ADT, some schedules for IR patients included ADT, and a majority of HR patients received ADT. For IR and HR patients, we also analyzed separately schedules that included ADT for most patients (≥50%) and those that did not: 9/32 IR and 15/20 HR schedules included ADT according to this definition.

An overview of the schedules included in the analysis is presented in Table 1, and further detailed information is presented in Supplementary Table 1.

**Table 1:** Overview of the characteristics of the schedules included in the analysis.

| Risk | Number of schedules | Number of patients (range) | Dose per fraction (range) | Total dose (range) | Treatment time (range) | ADT (fraction of schedules) | Control at 5 years (range) |
| --- | --- | --- | --- | --- | --- | --- | --- |
| LR | 35 | 3 – 550 | 1.8 – 10 Gy | 33.5 – 81 Gy | 3 – 62 days | 3/35 | 0.59 – 1.00 |
| IR | 32 | 7 – 839 | 1.8 – 10 Gy | 34 – 81 Gy | 3 – 62 days | 9/32 | 0.38 – 1.00 |
| HR | 20 | 12 – 821 | 1.8 – 8.5 Gy | 34 – 81 Gy | 3 – 62 days | 15/20 | 0.28 – 0.908 |

### Radiobiological modeling: dose-response

We have relied on the LQ-model to fit dose-response. The surviving fraction of tumor cells after a dose *d* is,

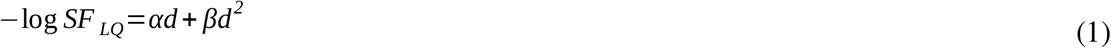

The LQL model **[18]**, which includes a moderation of the quadratic term of the LQ model with increasing dose, has also been investigated:

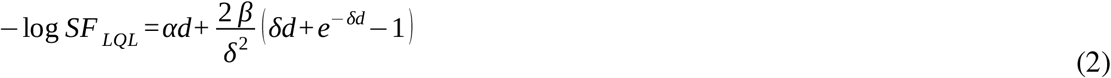

In addition, we have investigated an *ad hoc* modification of the LQ model presented in **[25]**, which includes an increasing effective quadratic term with increasing dose to account for indirect cell damage at large doses:

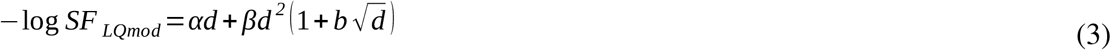

When delivering a treatment of *N* fractions, the overall surviving fraction is given by:

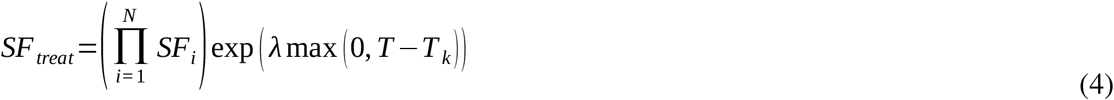

where *SF*_*i*_ is the surviving fraction associated to each fraction, *T* is the treatment time, and proliferation is modelled as exponential with rate *λ* after a kick-off time *T*_*k*_.

Tumour control probability was modelled using a logistic function **[28]**,

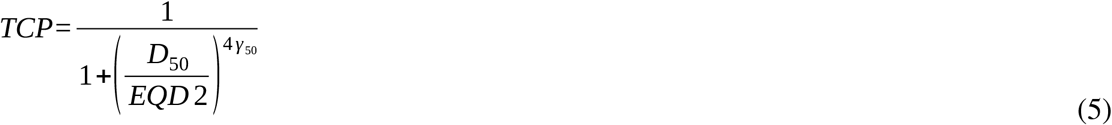

where *D*_50_, is the dose corresponding to 50% control (in 2 Gy fractions) and *γ*_50_ is the normalised dose-response gradient. *EQD2* is the equivalent dose in 2 Gy fractions of a given schedule, which is model dependent. For example, for the LQ model it can be calculated as:

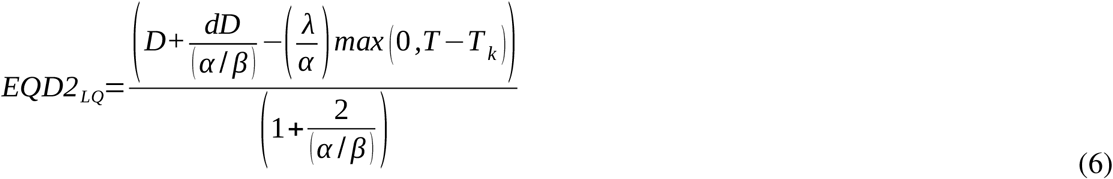

where *D, d* and *T* are the total dose, dose per fraction, and treatment time of the radiotherapy schedule. Similar equations can be written for the LQL and LQ_mod_ models using equations (2) and (3) to describe the effect of the *(d, D)* treatment and the equivalent 2 Gy treatment:

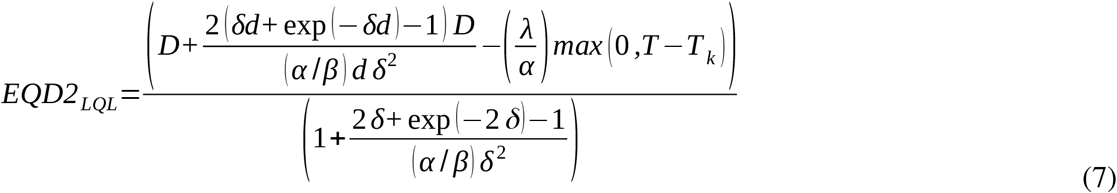

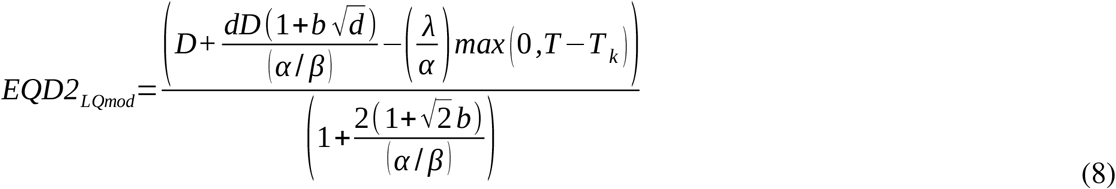

### Statistical methods

Fitting was performed by using the maximum likelihood methodology, assuming binomial statistics for the reported control values. The optimization (minimization of the -*log(L)* function, where *L* is the likelihood) was performed with an in-house developed simulated annealing algorithm.

The free parameters of the fit are *α/β, λ/α, T*_*k*_, *γ*_*50*_, and *D*_*50*_ for the LQ-model. For the LQL and LQ_mod_ models there is an extra parameter, *δ* and *b*, respectively. Notice that in this fit the value of *α* cannot be determined, only *α/β* (which conditions the response to different fractionation). The proliferation rate cannot be determined either, as it is entangled with *α*. We will define *λ’=λ/ α*, which has units of Gy/day, and it is related to the dose needed to compensate for repopulation.

The profile likelihood method was used to obtain 95% confidence intervals (CI) of best-fitting parameters **[29, 30]**. The implementation of the profile likelihood method is presented in more detail in the Supplementary Materials (Supplementary Figure 1).

The Akaike Information Criterion with sample size correction was used to rank different models **[31]**. The AIC_c_ is given by:

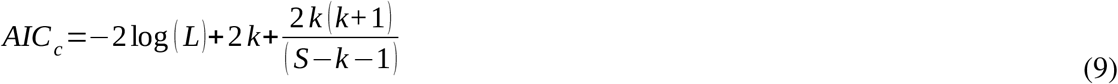

where *k* is the number of parameters of the model, *S* is the sample size, and *L* is the maximum of the likelihood function. Models with lower AIC_c_ are preferred. In this regard, the ΔAIC_c_ is defined as

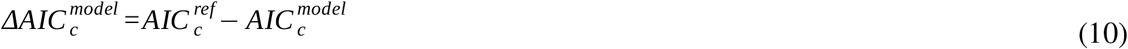

where AIC_c_^ref/model^ refers to the AIC_c_ of the reference model (the LQ model in this work) and the model under study.

The implementation of the methodology was performed in Matlab (Mathworks, Natick, MA).

### Radiobiological modeling: *α* and number of clonogens

Some further information on the radiobiology of the tumors can be obtained from the analysis of best-fitting parameters. Combining the TCP Poisson formulation **[32]** and the definition of D_50_ we can write (using the LQ model),

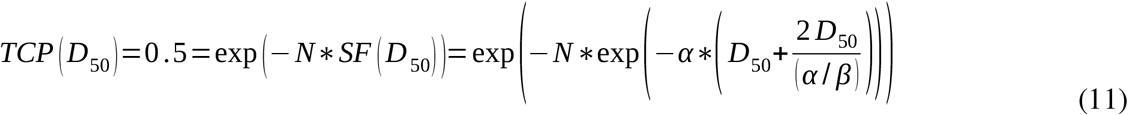

which can be converted to:

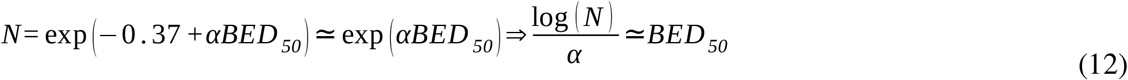

with

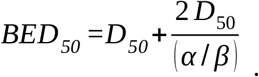

This expression provides a qualitative relationship between the number of clonogen cells (*N*), their radiosensitivity (*α*) and D_50_ (obtained from the fit to the dose-response data). Notice that for simplicity we have ignored the radiosensitivity averaging methodology that is usually included in the computation of TCP values with the Poisson model.

## Results

In Table 2 we present best-fitting parameters and goodness of fit (-log(L) and AIC_c_) obtained with the LQ, LQL and LQ_mod_ models for low, intermediate and high risk. For IR (HR) we also present separately fits for schedules that do not include ADT (include ADT).

**Table 2:** Best fits obtained with the LQ, LQL, and LQ_mod_ models to prostate carcinoma dose-response data, separated by risk (low, intermediate and high risk). For intermediate risk, results are also presented separately for schedules not including ADT. For high risk, results are also presented separately for schedules including ADT. The table shows best fitting parameters, maximum likelihood and AIC_c_ values. Improvements on the performance of the LQ model (ΔAIC_c_ >0) are highlighted in red. The symbol * indicates that the best-fitting parameter reached the edge of the constraint window.

| Risk | Model | Parameters |  |  |  |  |  |  |  |  |  |
| --- | --- | --- | --- | --- | --- | --- | --- | --- | --- | --- | --- |
| | | $\alpha/\beta$<br>[Gy] | $\lambda'$<br>[Gy day<sup>-1</sup>] | $T_k$<br>[day] | $\delta$<br>[Gy<sup>-1</sup>] | $b$<br>[Gy<sup>-1/2</sup>] | $D_{50}$<br>[Gy] | $\gamma_{50}$ | $-\log(L)$ | $AIC_c$ | $\Delta AIC_c$ |
| LR | LQ | 2.01 | 0.0 | - | - | - | 56.24 | 2.17 | 89.37 | 190.81 | - |
|  | LQL | 2.01 | 0.0 | - | 0.0 | - | 56.24 | 2.17 | 89.37 | 193.74 | -2.93 |
| | $LQ_{mod}$ | 2.60 | 0.0 | - | - | 0.07 | 55.78 | 2.11 | 89.19 | 193.38 | -2.57 |
| IR | LQ | 3.44 | 0.41 | 23.99 | - | - | 56.88 | 2.14 | 220.93 | 454.17 | - |
|  | LQL | 0.40 | 0.0 | - | 0.28 | - | 62.82 | 2.18 | 201.32 | 418.00 | 36.17 |
| | $LQ_{mod}$ | 3.46 | 0.41 | 23.94 | - | 0.0 | 56.84 | 2.15 | 220.93 | 457.22 | -3.05 |
| HR | LQ | 2.78 | 0.35 | 21.00* | - | - | 59.82 | 1.45 | 105.04 | 224.37 | - |
|  | LQL | 2.78 | 0.35 | 21.00* | 0.0 | - | 59.82 | 1.45 | 105.04 | 228.54 | -4.17 |
| | $LQ_{mod}$ | 11.16 | 0.34 | 21.00* | - | 0.75 | 58.67 | 1.47 | 103.86 | 226.18 | -1.81 |
| IR (no ADT) | LQ | 2.83 | 0.32 | 21.00* | - | - | 58.10 | 1.85 | 157.58 | 328.69 | - |
|  | LQL | 0.46 | 0.0 | - | 0.24 | - | 63.63 | 2.01 | 138.66 | 294.57 | 34.12 |
| | $LQ_{mod}$ | 2.83 | 0.32 | 21.00* | - | 0.0 | 58.10 | 1.85 | 157.58 | 332.41 | -3.72 |
| HR (ADT) | LQ | 2.09 | 0.0 | - | - | - | 58.46 | 0.95 | 72.55 | 161.77 | - |
|  | LQL | 2.08 | 0.0 | - | 0.0 | - | 58.45 | 0.95 | 72.54 | 167.58 | -5.81 |
| | $LQ_{mod}$ | 18.71 | 0.0 | - | - | 1.99 | 56.75 | 0.88 | 71.19 | 164.88 | -3.11 |

For LR and HR best-fits obtained with the LQL model have *δ*≈*0*, and therefore best-fitting solutions are almost identical to those obtained with the LQ model. Because the LQL has one extra degree of freedom, this results in higher AIC_c_ than those obtained with the LQ (ΔAIC_c_<0). For IR, the LQL clearly outperforms the LQ (and LQ_mod_) model, with ΔAIC_c_≈36 (Table 2 and Supplementary Figure 2). On the other hand, best-fits obtained with the LQ_mod_ show a very modest improvement over the LQ model for LR and HR when comparing -log(L), but due to the extra parameter it does not lead to ΔAIC_c_ >0.

Best-fitting parameters obtained with the LQ model are presented in more detail in Table 3, including 95% CI. Results for intermediate and high risk are also presented separately for cohorts including/not including ADT as part of treatment. Best-fits show low *α/β* values (2.01 Gy for LR, 3.44 Gy for IR and 2.78 Gy for HR), while 95% CI are [1.74-2.26] Gy for LR, [2.99-4.02] Gy for IR and [1.43-4.18] Gy for HR. D_50_ values range from 56.24 Gy for LR to 59.82 Gy for HR. The results show no proliferation for LR tumors, and proliferation rates (kick-off time) values of 0.41 Gy/day (24 days) for IR, and 0.35 Gy/day (21 days) for HR. It is important to notice that we implemented a minimum constraint of 21 days for *T*_*k*_.

**Table 3:** Best-fitting parameters and 95% confidence intervals (within parentheses) of prostate carcinoma dose response data obtained with the linear quadratic model. Results are separated by risk, and for intermediate and high risk are also presented separately for schedules that include or do not include ADT. Data for IR with ADT and HR with no ADT are shown only for illustrative purpose, because due to the low number of schedules confidence intervals are very wide.

| | $\alpha/\beta$ [Gy] | $\lambda'$ [Gy day<sup>-1</sup>] | $T_k$ [day] | $D_{50}$ [Gy] | $\gamma_{50}$ |
| --- | --- | --- | --- | --- | --- |
| **Low Risk** | 2.01<br>(1.74, 2.26) | 0<br>(0, 0.13) | - | 56.24<br>(54.36, 57.99) | 2.17<br>(1.90, 2.47) |
| **Intermediate Risk** | 3.44<br>(2.99, 4.02) | 0.41<br>(0.31, 0.49) | 23.99<br>(21.0, 25.5) | 56.88<br>(55.51, 57.91) | 2.14<br>(1.92, 2.40) |
| **High Risk** | 2.78<br>(1.43, 4.18) | 0.35<br>(0, ∞) | 21.0<br>(21.0, ∞) | 59.82<br>(57.07, 63.93) | 1.45<br>(1.07, 1.83) |
| **Intermediate Risk**<br>**(no ADT)** | 2.83<br>(2.10, 3.47) | 0.32<br>(0.09, 0.46) | 21.0<br>(21.0, 27.3) | 58.10<br>(56.51, 60.04) | 1.85<br>(1.55, 2.14) |
| **High Risk (ADT)** | 2.09<br>(1.50, 3.46) | 0<br>(0, 0.31) | - | 58.46<br>(54.34, 61.54) | 0.95<br>(0.75, 1.25) |
| **Intermediate Risk**<br>**(ADT)** | 0.1<br>(0, ∞) | 0<br>(0, ∞) | - | 8.10<br>(0.43, 40.50) | 0.20<br>(0.11, 0.80) |
| **High Risk (no ADT)** | 100.00<br>(7.08, ∞) | 3.31<br>(1.09, ∞) | 39.9<br>(21.0, 40.8) | 54.58<br>(49.75, 60.49) | 6.68<br>(2.70, 10.67) |

When analyzing separately data for IR/HR patients that were treated with ADT or not, we obtained *α/β* =2.83 Gy, D_50_ = 58.10 Gy, *λ*’=0.32 Gy/day, *T*_*k*_ = 21 days for IR “only RT”, and *α/β* =2.09 Gy, D_50_ = 58.46 Gy, and no proliferation for HR “RT+ADT”. Best fits for IR “RT+ADT” and HR “only RT” are also presented in Table 3, but due to the low number of schedules involved (9 and 5 respectively) confidence intervals are very wide.

In Figure 1 we show best-fits to prostate carcinoma dose-response data obtained with the linear quadratic model. Results are presented separately for LR, IR and HR. In Figure 2, best-fits for IR and HR are shown separately for cohorts including ADT and cohorts not using ADT in addition to radiotherapy.

**Figure 1:**
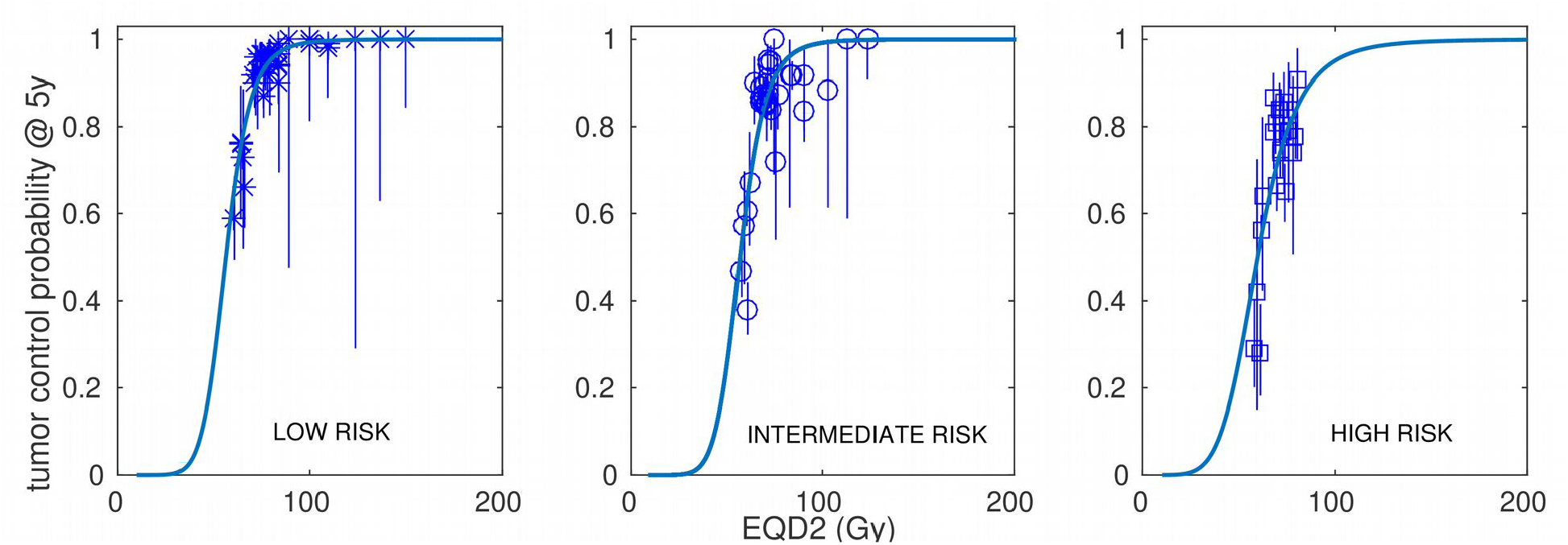
Best fits to prostate carcinoma dose-response data obtained with the linear quadratic model. Results are presented separately for low risk (left panel), intermediate risk (central panel) and high risk (right panel).

**Figure 2:**
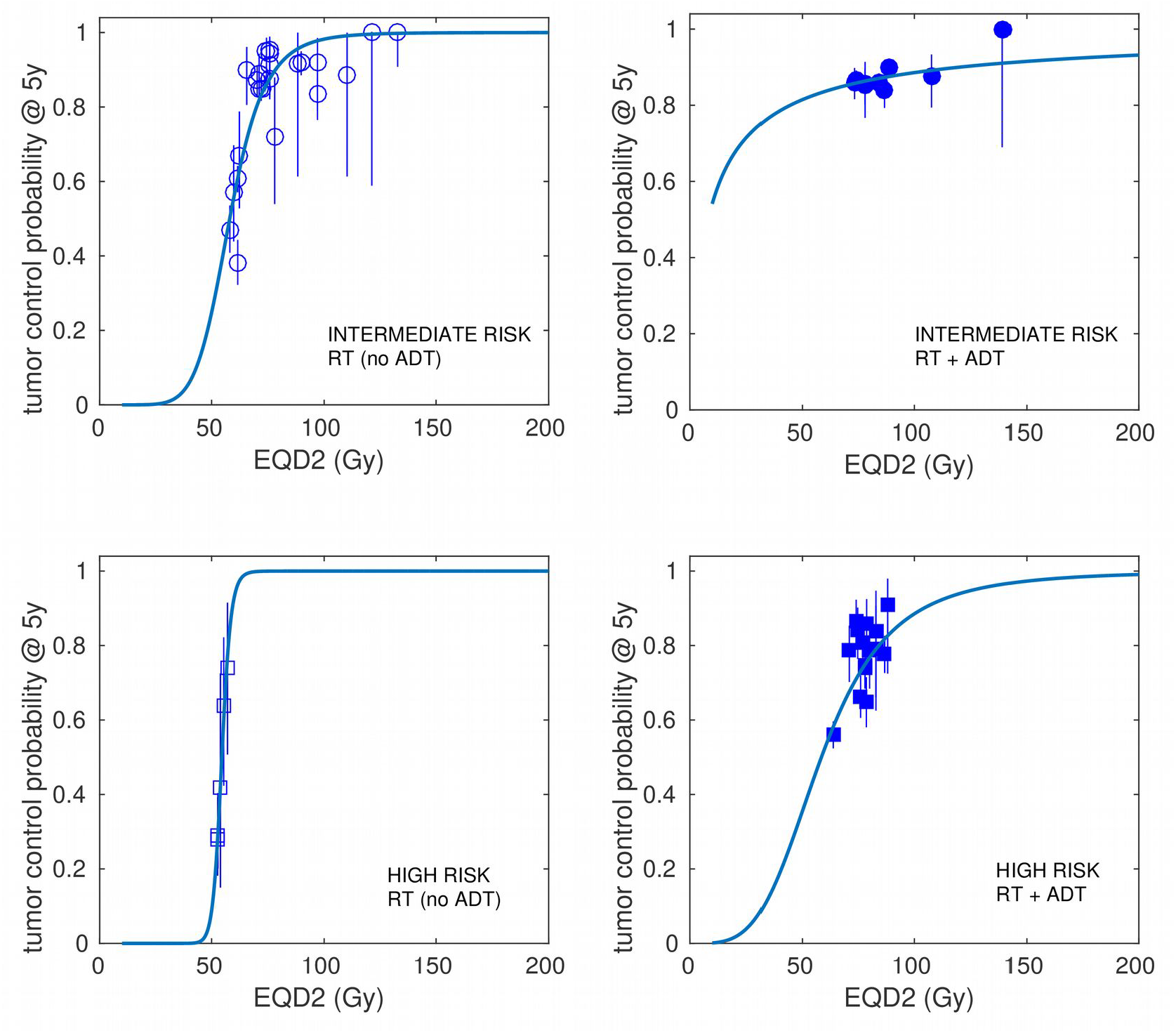
Best fits to intermediate- and high-risk prostate carcinoma dose-response data obtained with the linear quadratic model. Results are presented separately for cohorts that use androgen deprivation therapy (ADT) and cohorts not using ADT in addition to radiotherapy.

We have investigated the dose per fraction versus number of fractions that would be necessary to obtain 90% for HR patients treated with radiotherapy and ADT according to best-fitting parameters obtained with the LQ model. These results are presented in Figure 3, where we also present the experimental fractionations included in the dataset for “RT+ADT”.

**Figure 3:**
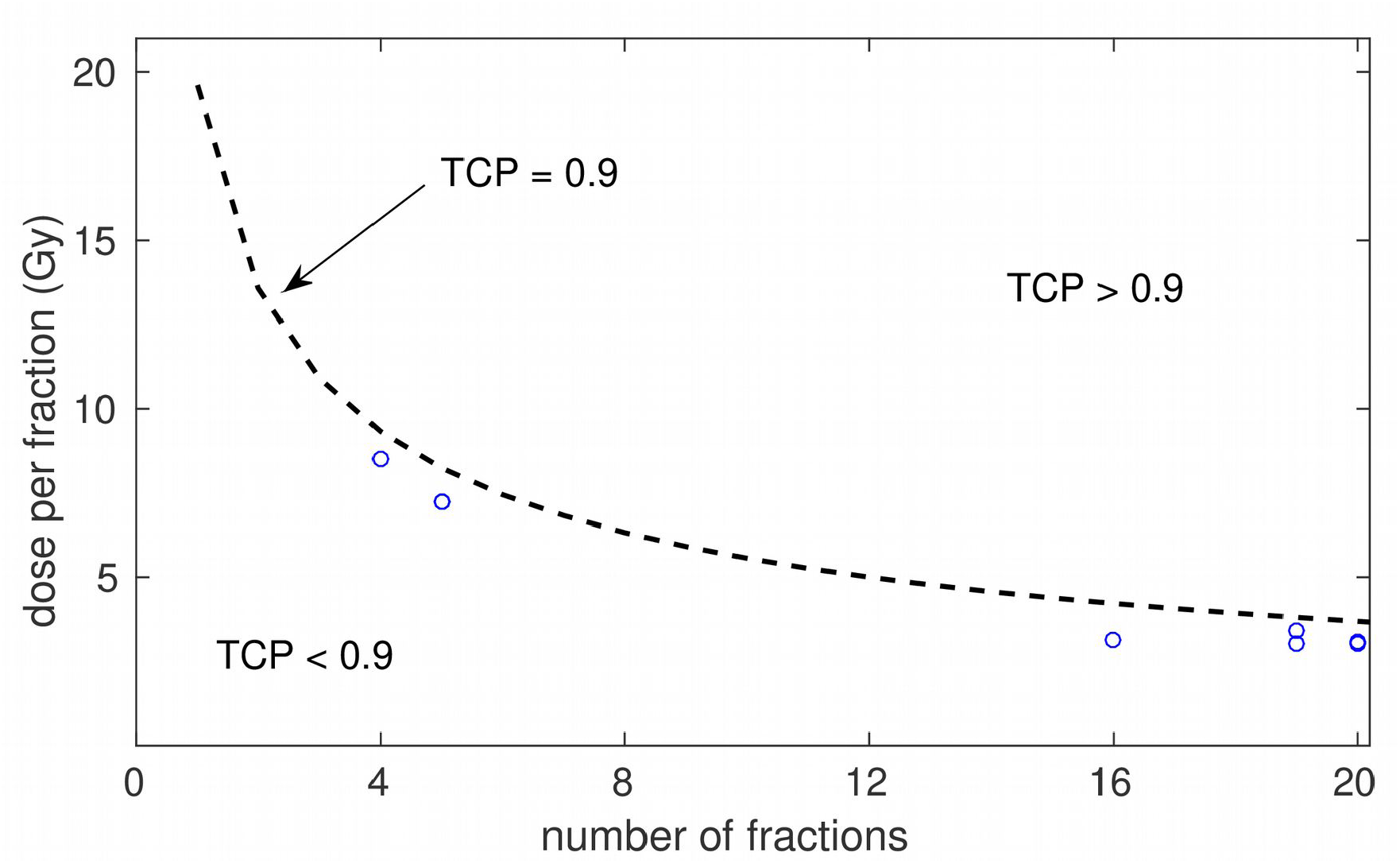
Modelled dose per fraction versus number of fractions to achieve 90% control for HR patients treated with radiotherapy and ADT (dashed line). The circles represent the experimental fractionations included in the dataset

Applying equations (11) and (12), which qualitatively link the number of clonogens and radiosensitivity of tumor cells, to the best-fitting parameters obtained with the LQ model, we obtain:

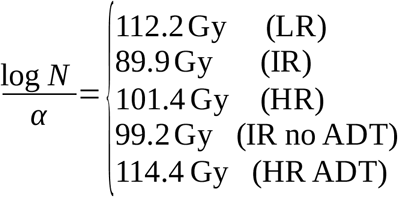

If we assume N_HR_ > N_IR_ > N_LR_ (which is supported by the analysis of Pedicini et al **[9]**, who reported N_LR_=4.5e5, N_IR_=3e6, N_HR_=2e7), we may conclude that LR cells might be less radiosensitive than HR/IR cells (by using the numbers of cells reported in **[9]** we obtain *α*_*LR*_∼0.12 Gy^-1^, versus *α*_*IR/HR*_∼0.17 Gy^-1^).

## Discussion

It has been suggested that the LQ model may fail to describe dose-response at large doses per fraction due to the contribution of effects like damage repair, vascular damage or radiation-induced immune effects **[15-17]**. Therefore, we have investigated not only the LQ model, but also other models that include departures from the LQ behavior at large doses per fraction (the LQL model, with decreasing radiosensitivity with increasing dose, and a phenomenological modification of the LQ model with increasing radiosensitivity with increasing dose).

Fits with the LQ_mOd_ show a very modest improvement over the LQ model for LR and HR (ΔAIC_c_ ≈ 0.1). Analyses based on the AIC typically set stronger thresholds, demanding ΔAIC_c_>6 to state the superiority of a given model over another **[33]**. On the other hand, fits with the LQL model show a clear improvement over the LQ for IR patients (ΔAIC_c_ > 30).

The superiority of the LQL over the LQ for IR merits further discussion. An analysis of the schedules included in the dataset shows that the superiority of the LQL is strongly conditioned by a schedule reported in a recent study by Levin-Epstein *et al*. **[34]**. In that work, they reported control for 1904 patients treated with SBRT, including 157 intermediate risk patients (93 favorable, 64 unfavorable) treated with 38 Gy in 4 fractions (9.5 Gy per fraction). Control at 5 years for those patients was 83.6% (86.7% for favorable and 79.2% for unfavorable), well below control obtained in the same risk group for 35Gy/5f (89.0%), 36.25Gy/5f (95.2%) and 40Gy/5f (92.0%). If we exclude the 38Gy/4f results from the analysis, the ΔAIC_c_ for the LQL decreases from 36 to 6.

In our dataset there are schedules delivering similar doses-per-fraction that report higher controls, but they include a much lower number of patients *(e*.*g*. 38Gy/4f, control=92%, 39 patients). The relative low control rates obtained for a dose per fraction of 9.5 Gy may be a hint of LQL behavior at large doses, but should be confirmed by more experimental studies.

In addition, the fact that the superiority of the LQL model is observed only for IR may be related to the poor goodness of fit obtained for IR (-log(L)>200 vs -log(L)≈100 for LR/HR). The worse fits obtained for IR could be caused by a more heteregeneous dataset (caused by different ratios of favorable/unfavorable IR patients or more heterogeneity in the administration of ADT).

We cannot discard that other models may provide a better fit to the experimental data. For example, models accounting for hypoxia and reoxygenation, which have been suggested that play a role in the response of prostate cancer **[8, 35]**, have not been investigated. In this regard, the large dataset that we have assembled (Supplementary Table 1) may prove useful for other researchers to investigate different models.

The analysis based on the LQ model supports a low *α/β* value for all risk groups of prostate cancer, with 95% CI of [1.74-2.26] Gy for LR, [2.99-4.02] Gy for IR and [1.43-4.18] Gy for HR. Nonetheless, our analysis shows that the *α/β* of IR is larger than that of LR, which may be taken into account when designing optimal fractionations. The low *α/β* values are in general agreement with several radiobiological analyses of dose-response in prostate cancer **[1-7, 9-10]**. However, most of these studies did not include hypofractionated treatments (only **[10, 27]**) and/or analyzed a lower number of schedules.

High risk, and to less extent intermediate risk, prostate cancer is usually treated with a combination of radiotherapy and ADT. When analyzing separately HR cohorts including ADT or not, it seems that the addition of ADT eliminates tumor proliferation (*λ’*=0 Gy/day for HR cohorts including ADT versus *λ’*=0.39 Gy for all HR cohorts). It would be of interest to know whether the addition of ADT affects the *α/β* of the tumor. However, due to the low number of HR schedules that do not include ADT (and IR schedules that include ADT), confidence intervals are very wide, and no conclusive evidence can be reported on differences between adding ADT or not.

Control rates for LR and IR prostate cancer are typically above 90%. However, control rates for HR prostate cancer are lower. We have investigated the dose per fraction that is necessary to obtain 90% for HR patients treated with radiotherapy and ADT according to best-fitting parameters obtained with the LQ model. Experimental schedules included in the dataset are below the TCP=0.9 boundary (see Figure 3). According to the model, doses per fraction of 10.9 Gy, 8.2 Gy and 5.6 Gy are needed to reach 90% control with 3, 5 and 10 fractions. Whether the toxicity associated to such dose escalation is tolerable has not been studied in this work. It may be worth exploring hypofractionated dose escalation schedules aiming at increasing the control rate of HR cancer for subsets of patients who are genetically less predisposed to suffer toxicity **[36]**.

In conclusion, the analysis of dose-response of prostate cancer did not show evidence of effects beyond the LQ contributing at large doses per fraction, except for IR schedules where the LQL is superior to the LQ, pointing out a possible moderation of radiosensitivity with increasing dose. Our analysis shows a low *α/β* for all risks of prostate cancer. However, the *α/β* for IR (95% CI [2.99-4.02] Gy) is significantly larger than for LR (95% CI [1.74-2.26] Gy).

## Data Availability

All data produced in the present work are contained in the manuscript and supplementary materials.

## Acknowledgements

This project has received funding from Ministerio de Ciencia e Innovación, Agencia Estatal de Investigación and FEDER, UE (grant PID2021-128984OB-I00). This project has received funding from Xunta de Galicia, Axencia Galega de Innovación (grant IN607D 2022/02).

## Supplementary Materials

### 1. Statistical methods

The profile likelihood method was used to obtain confidence intervals of best-fitting parameters. A parameter value *θ*_*0*_ will be in the (1-α) confidence interval if it verifies

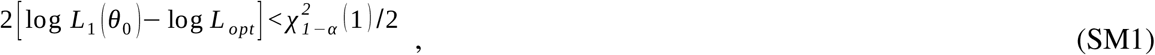

where log *L*_*opt*_ is the likelihood of the full model, log *L*_1_(*θ*_*0*_) is the likelihood of the model when the value of the parameter *θ* is set to *θ*_*0*_, and χ^2^_1−α_ (1) is the (1−α) quantile of a χ^2^ distribution with 1 d.f.

In order to obtain confidence intervals for a given parameter *θ*, we evaluated equation (SM1) for a set of { *θ*_*0*_} values, typically 8-12, paying attention to having values off the 95% confidence interval on both the left and right side. The set of {*θ*_*0*_, log *L*_1_(*θ*_*0*_)-log *L*_*opt*_ } were interpolated with a shape-preserving piecewise cubic hermite interpolating polynomial (pchip), and the specific values defining the left (*θ*_L_) and right (*θ*_R_) sides of the confidence interval, verifying

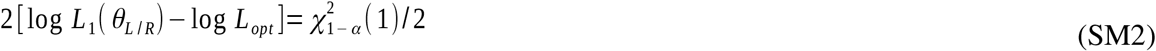

were obtained from the interpolation.

The profile likelihood method was implemented with one particularity: If best-fitting parameters showed no proliferation, the confidence interval of the proliferation rate *λ’* was obtained by fixing *T*_*k*_ = 21 days (the minimum kick-off time allowed in this study). Fixing the value of *T*_*k*_ is necessary, otherwise the optimizer will obtain optimal non-proliferative solutions, equivalent to *λ’*=0, by selecting large values of *T*_*k*_ (and therefore 2[log L_1_(*λ*’) -log L_opt_]=0 for all *λ’*).

In Supplementary Figure 1 we illustrate this calculation for the *α/β* parameter in IR.

### 2. Supplementary Figures

**Supplementary Figure 1:**
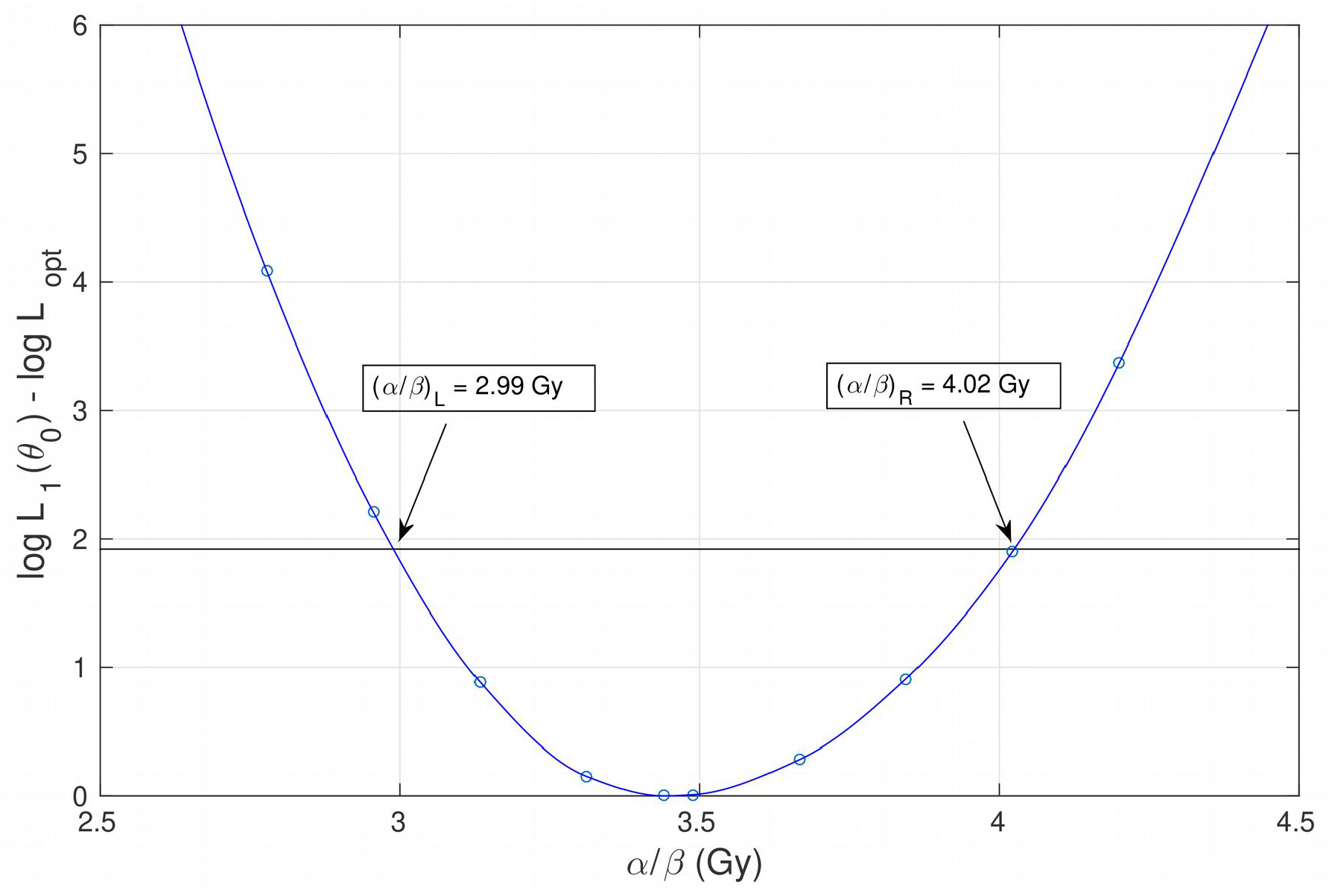
Illustration of the calculation of 95% confidence intervals for the parameter (α/β) in IR patients.

**Supplementary Figure 2:**
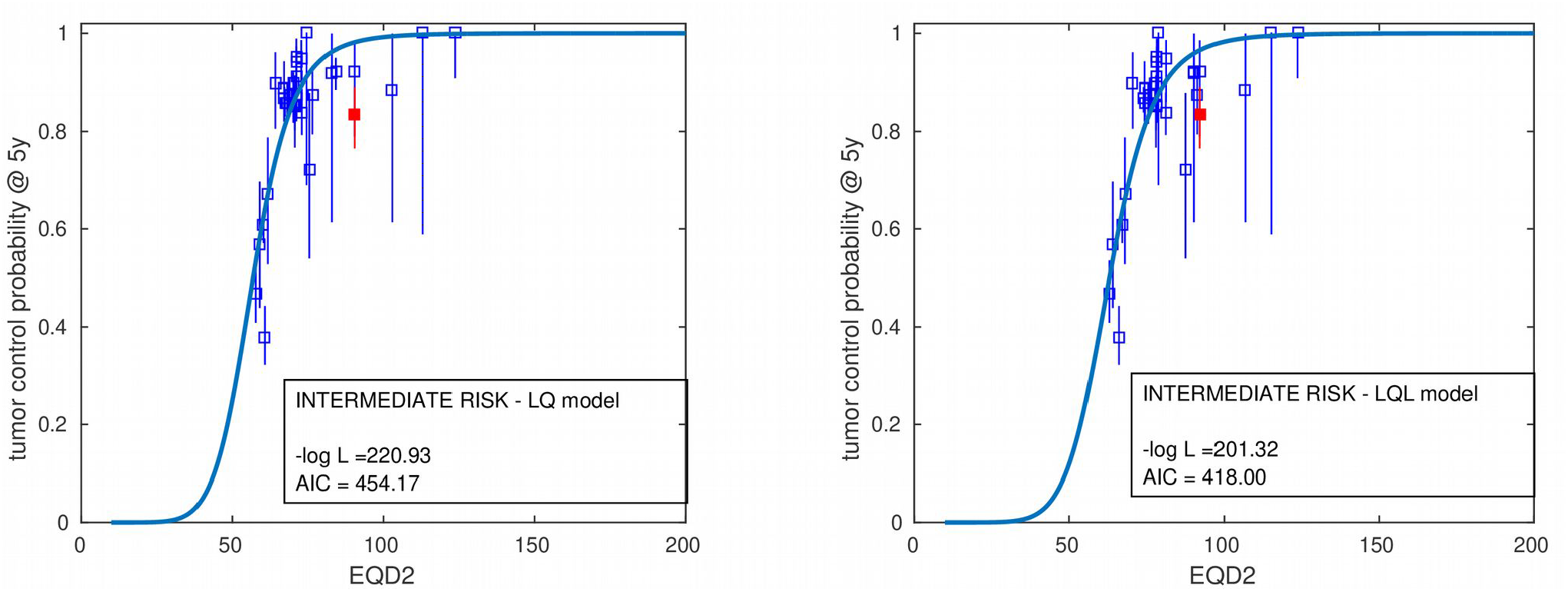
Best fits to intermediate risk dose-response data for prostate cancer obtained with the linear quadratic (LQ) and linear-quadratic-linear (LQL) models: left panel and right panel, respectively. In red we highlight the schedule that conditions the superiority of the LQL model: reported by Levin-Epstein *et al*., it includes 157 intermediate risk patients treated with 38 Gy in 4 fractions (9.5 Gy per fraction), and control at 5 years was 83.6%

### 3. Supplementary Tables

**Supplementary Table 1:**
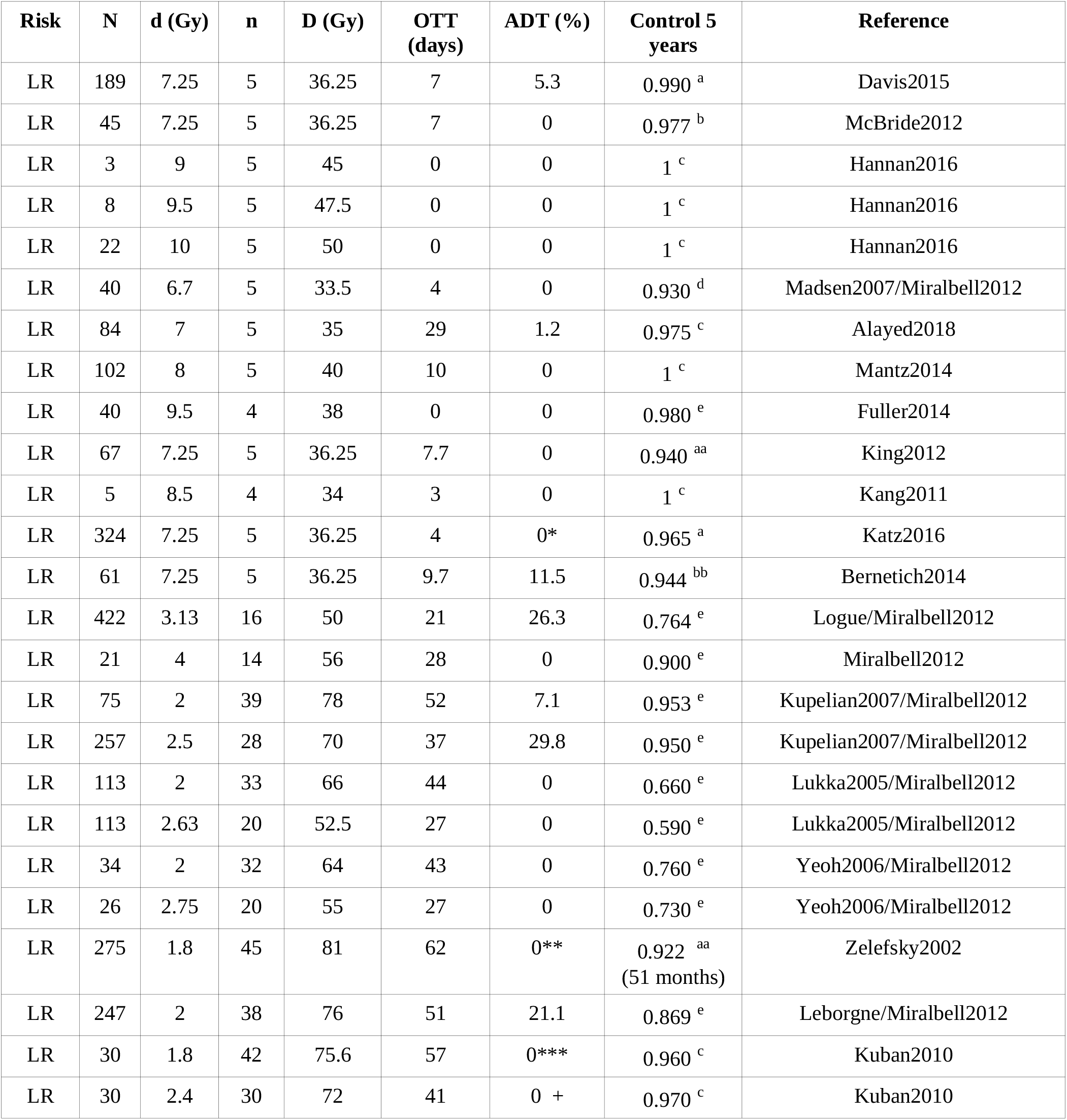
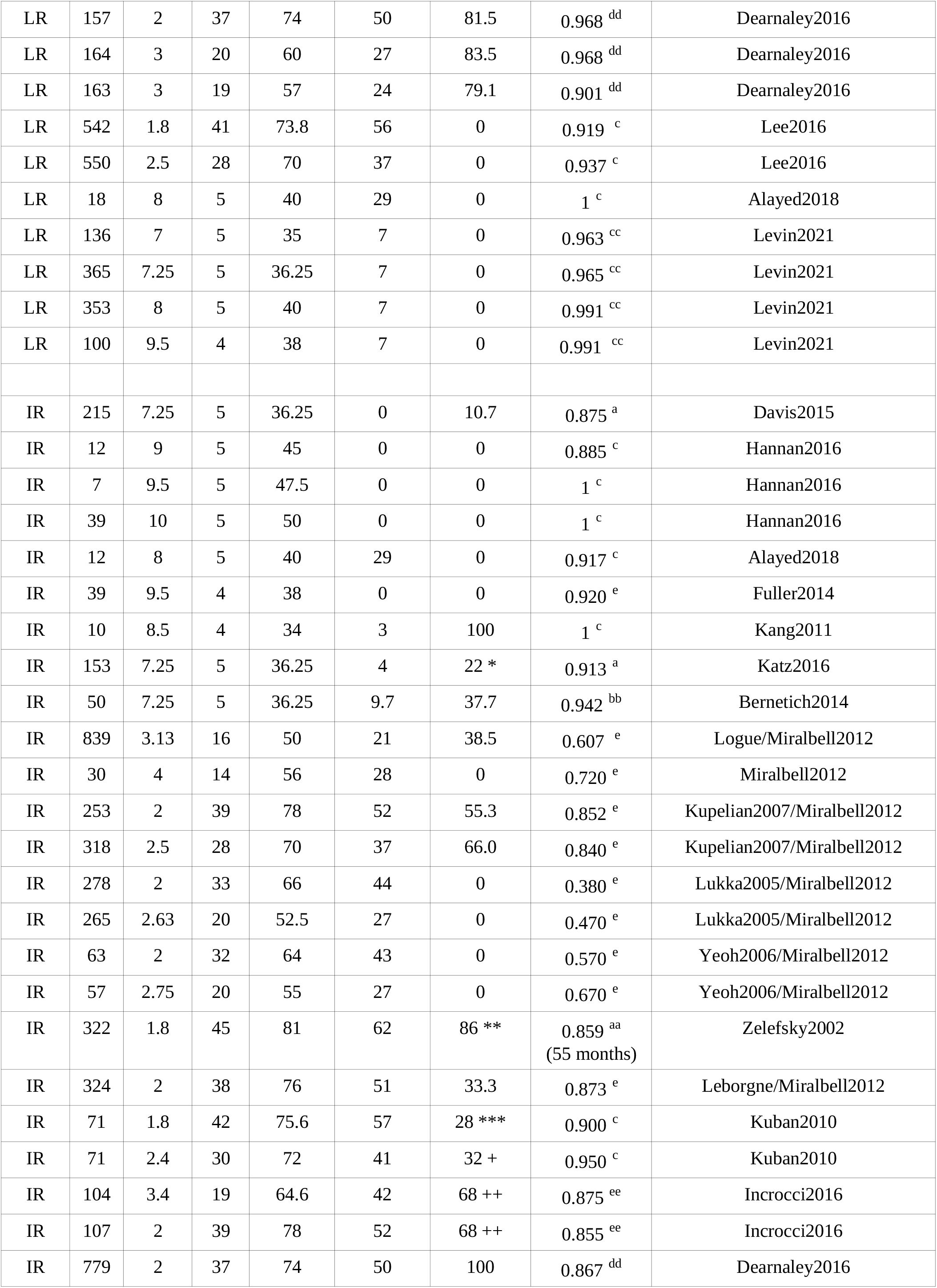
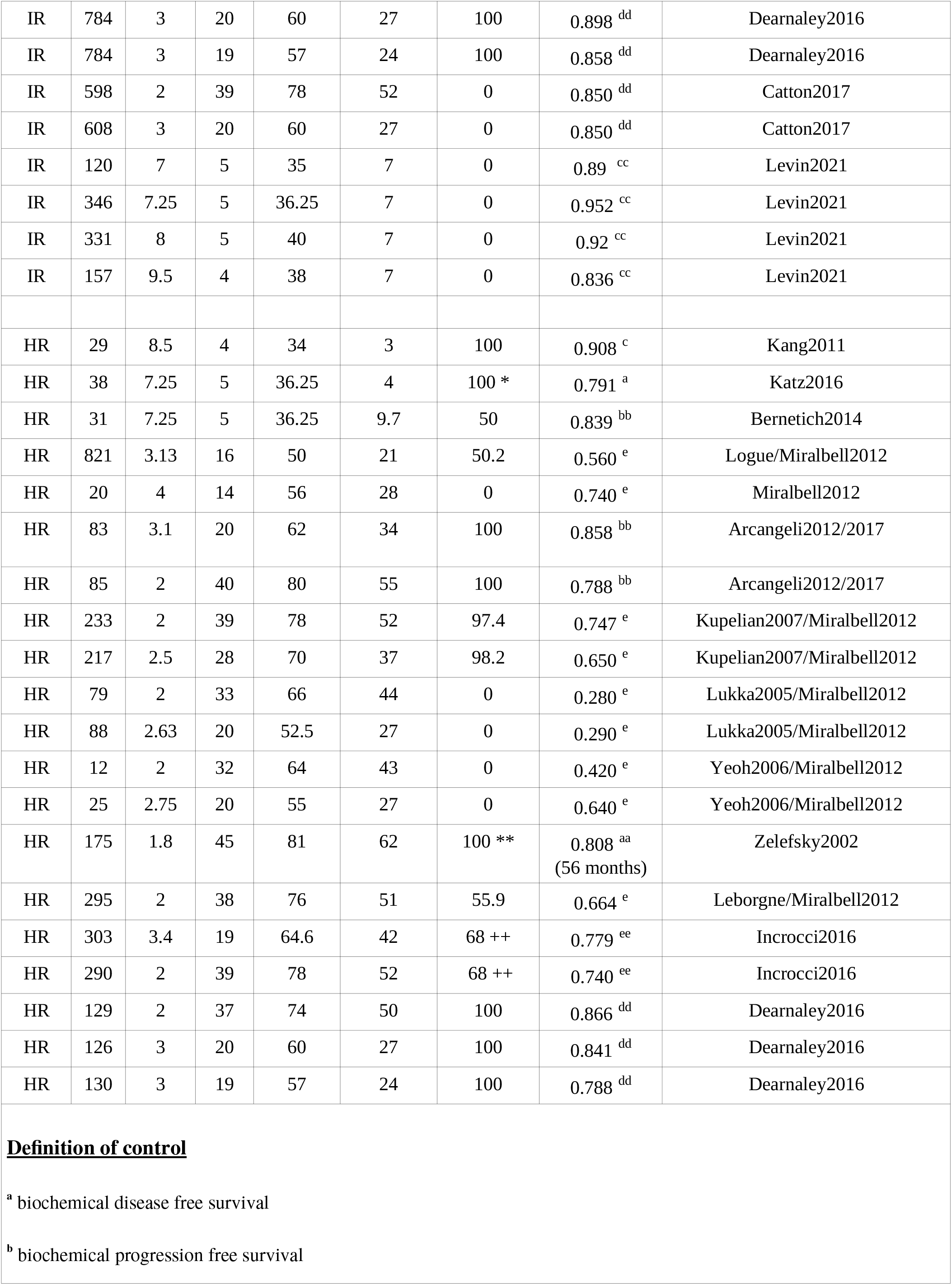
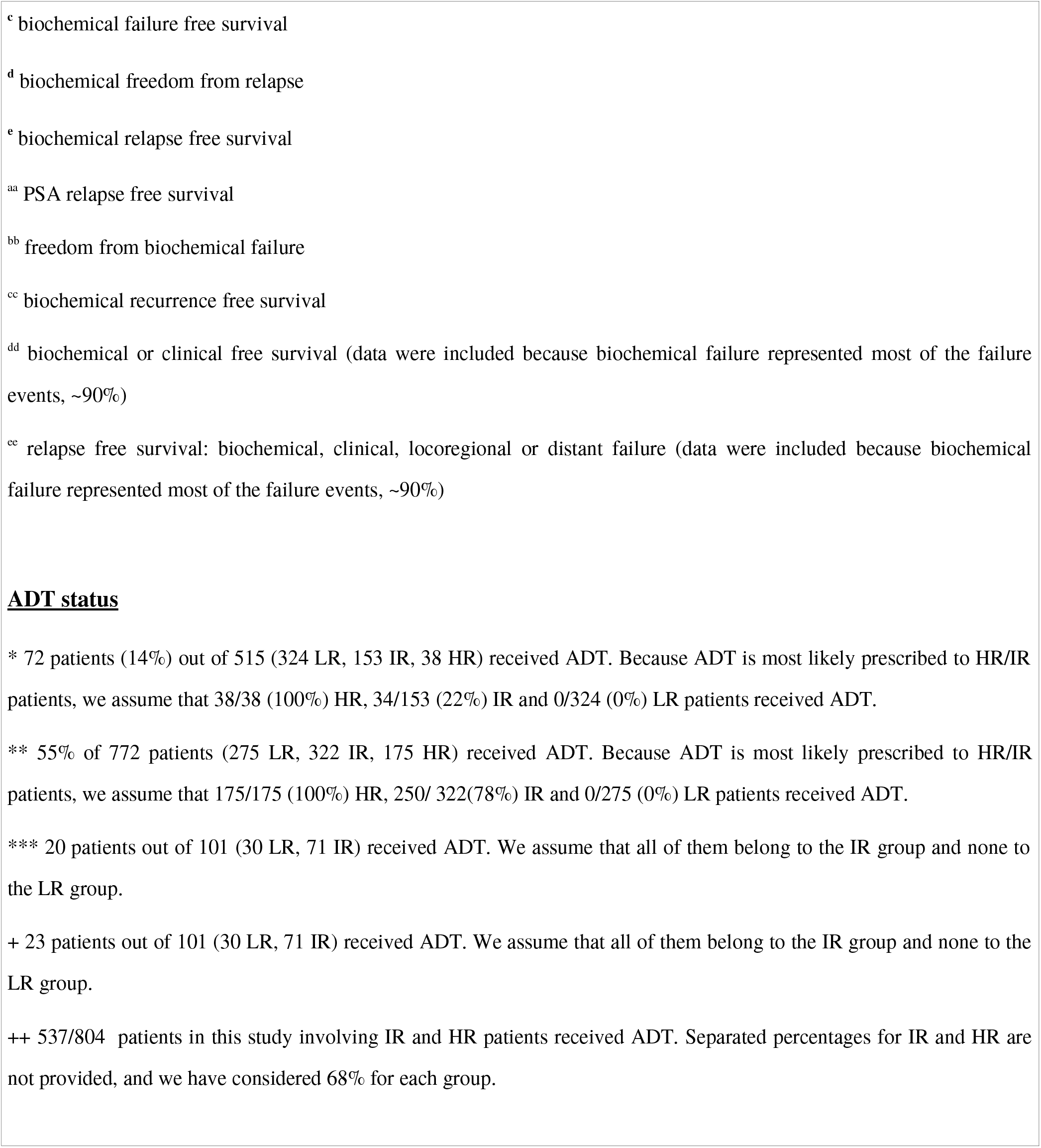
Detailed information of the analyzed schedules for low (LR), intermediate (IR), and high risk (HR) prostate cancer, including: number of patients (N), dose per fraction (d), number of fractions (n), total dose (D), overall treatment time (OTT), percentage of patients receiving ADT, control at five years, and the first author and year of the study.

